# Associations between life course longitudinal growth and hip shapes at ages 60 to 64 years: evidence from the MRC National Survey of Health and Development

**DOI:** 10.1101/2023.10.12.23296966

**Authors:** Katherine A. Staines, Fiona R. Saunders, Alex Ireland, Richard M. Aspden, Jenny S. Gregory, Rebecca Hardy, Rachel Cooper

## Abstract

We sought to examine associations between height gain across childhood and adolescence with hip shape in individuals aged 60-64 years from the MRC National Survey of Health and Development, a nationally representative British birth cohort. Height was measured at ages 2, 4, 6, 7, 11 and 15 years, and self-reported at age 20 years. Ten modes of variation in hip shape (HM1-10), described by statistical shape models, were previously ascertained from DXA images taken at ages 60-64 years. Associations between (1) height at each age (2) Super-Imposition by Translation And Rotation (SITAR) growth curve variables of height size, tempo and velocity, and (3) height gain during specific periods of childhood and adolescence, and HM1-10 were tested. Faster growth velocity was associated with a wider, flatter femoral head and neck, as described by positive scores for HM6 (regression coefficient 0.014; 95%CI, 0.08–0.019; P<0.001) and HM7 (regression coefficient 0.07; 95%CI, 0.002-0.013; P=0.009), and negative scores for HM10 (regression coefficient –0.006; 95%CI, – 0.011-0.00, P=0.04) and HM2 (males only, regression coefficient –0.017; 95%CI, –0.026 – – 0.09; P<0.001). Similar associations were observed with greater height size and later height tempo. Examination of height gains during specific periods of childhood and adolescence identified those during the adolescence period as being most consistently associated. Our analyses suggest that individual growth patterns, particularly in the adolescent period, are associated with modest variations in hip shape at 60-64 years, which are consistent with features seen in osteoarthritis.

## Introduction

Osteoarthritis, characterised by articular cartilage loss, subchondral bone thickening and osteophyte formation, is a global health care burden affecting about 528 million people worldwide in 2019 [1]. There are currently no disease modifying treatments available, and once disease has advanced, total joint replacement is the only option. Therefore, a better understanding of the mechanisms underpinning osteoarthritis, and in particular identification of predictors of disease risk, could help inform strategies for prevention and treatment.

Joint shape is known to be closely related to osteoarthritis predisposition. Indeed, developmental dysplasia of the hip is a known risk factor for both human and animal osteoarthritis [2]. Similarly, we and others have shown in numerous studies using statistical shape modelling (SSM), a sophisticated technique that enables identification of subtle shape variations, that hip shape is associated with radiographic osteoarthritis [3–5]. The most likely mechanism by which hip shape may predict osteoarthritis predisposition is through a biomechanical influence on joint loading, however there may be alternative genetic and molecular mechanisms contributing to this relationship. Indeed, several known osteoarthritis susceptibility single nucleotide polymorphisms (SNPs) have been associated with hip shape in perimenopausal women in the Avon Longitudinal Study of Parents and Children (ALSPAC) [6]. Most intriguingly, when combined with data from other cohorts, the eight SNPs independently associated with hip shape were intimately associated with the process of endochondral ossification [7].

Concurrent with this, a study from offspring in the ALSPAC found height tempo (corresponding to pubertal timing) to be strongly associated with the hip shape modes which may be related to future risk of hip osteoarthritis [8]. Moreover, we have examined associations between lifetime linear growth trajectories using the SuperImposition by Translation and Rotation (SITAR) variables of height size, tempo and velocity, and knee osteoarthritis in the MRC National Survey of Health and Development (NSHD). We found that increased height in childhood was associated, albeit modestly, with lower odds of knee osteoarthritis at age 53 years, as was adult achieved height [9]. Adult achieved height has also been shown to correlate with subtle variations in hip shape in this cohort [4]. Similarly, age at onset of walking is associated with hip shape in early old age in this cohort, and these associations were not explained by adjustment for lean mass, suggesting that early life skeletal loading may play a key role in the underlying mechanism [10]. Whether linear growth and in particular, its timing and velocity, during childhood and adolescence are associated with hip shape and osteoarthritis development is unknown.

This has therefore led us to hypothesise that life course longitudinal growth may be associated with variations in hip shape relevant to the development of osteoarthritis, which has important implications for musculoskeletal health in aged individuals. Herein, we interrogated the MRC NSHD to examine the relationships between height gain across childhood and adolescence with hip shape in individuals aged 60 to 64 years. Our aims were to test associations of: (1) height during childhood and adolescence; (2) SITAR parameters of height size, tempo, and velocity; (3) height gain during specific life periods, with hip shapes characterised using SSM of DXA images taken at age 60-64 years.

## Materials and Methods

### Study sample

The MRC NSHD is a birth cohort study, which includes a nationally representative sample of 2815 men and 2547 women born in England, Scotland, and Wales during 1 week in March 1946 [11,12]. The cohort has been followed prospectively across life with outcome data for these analyses drawn from a data collection between 2006 and 2010. Between these dates, eligible participants known to be alive and living in England, Scotland, and Wales were invited for an assessment at one of six clinical research facilities. Of 2,856 individuals invited, 1690 attended a clinical research facility and 539 received a home visit from a research nurse. Ethical approval for this data collection was obtained from the Central Manchester Research Ethics Committee (07/H1008/245) and the Scottish A Research Ethics Committee (08/MRE00/12) and participants provided written informed consent.

### Hip DXA images & statistical shape modelling

During their visit to a clinical research facility at age 60-64, images of the participant’s total body and left hip (except in 63 cases where contraindication of a prosthesis meant that the right hip was scanned) were obtained using a QDR 4500 Discovery DXA scanner (Hologic, Inc., Bedford, MA, USA). All hip scans were performed with the feet placed at 15 degrees of internal rotation. In five centres, scanners had rotating CLJarms allowing participants to lie supine for all scans; one centre used a scanner with a fixed CLJarm. Of the 1690 participants who attended a clinical research facility, 1636 had a hip DXA scan. Three images were excluded because of extreme internal rotation of the femur, evidenced by femoral neck foreshortening, leaving 1633 images to build the hip SSM, as previously described [4]. Hip modes (HM) were identified which were standardized to a mean of 0 and SD of 1; each accounted for >2% hip shape variation. In total, these 10 modes accounted for 80.6% of the total variance.

### Height

Height (cm) was measured by nurses using standardised protocols at ages 2, 4, 7, 11, and 15 years, and self-reported at age 20 years. Individual patterns of height growth during puberty were estimated using the SITAR model of growth curve analysis, as previously described by Cole et al. [13,14] The SITAR model estimates the mean growth curve and three individual-specific parameters: size (reflecting differences in mean height), tempo (reflecting differences in the timing of the pubertal growth spurt) and velocity (reflecting differences in the duration of the growth spurt), each expressed relative to the mean curve.

### Covariates

Factors that may potentially confound the main associations of interest were selected *a priori* based on previous findings in the literature [9,15]. These were birth weight, father’s occupational class in childhood (categorised as non-manual vs manual) and sporting ability at 13 years (categorised as above average, average, or below average according to teacher reports of their sporting ability) [16,17]. Weight was measured by nurses using standardised protocols at ages 2, 4, 7, 11, and 15 years, and self-reported at age 20 years.

### Statistical analysis

We used linear regression models to test associations between: (1) height at each age (2) SITAR height variables and (3) conditional changes in height during specific periods of early life (early childhood: 2–4 years; late childhood: 4-7 years; childhood to adolescence: 7–15 years; adolescence to young adulthood: 15–20 years) and each hip shape mode.

In models to address (3), we generated conditional changes in height by regressing each height measure on the earlier height measure for each sex and calculating the residuals [18]. The residuals were standardized (to have mean 0 and SD of 1) to ensure their comparability and these were included as the main independent variables.

In initial models, we formally tested for interactions between sex and each main independent variable and where no evidence of interaction was found based on statistical significance (P<0.05), models were fitted with men and women combined and adjusted for sex. We also tested for deviations from linearity by including quadratic terms in all growth variables. In each set of models, we first adjusted for sex (where there was no evidence of interaction), before then also adjusting for early life factors (birth weight + sporting ability at 13 years + father’s occupational class in childhood). In our final model, we adjusted SITAR height for SITAR weight variables, and conditional height gain for conditional weight gain to assess the contribution of weight during growth. To maximise sample size when examining height at each age, each set of models were run on the sample with valid data for the height at that age, the HM and the covariates. For the SITAR analysis, models were run on the sample with valid data for SITAR variables, HM and the covariates. For the analysis of height gains during specific periods, models were run on the sample with valid data for height at all ages, HM and the covariates to ensure the same N to aid formal comparison of effect sizes across time periods. Data were analysed using Stata statistical software (version SE 14.2).

## Results

Characteristics of the participants included in this study are detailed in Table 1. A total of 770 men and 842 women had complete data on the SITAR parameters of height, and the hip shape modes. In this sample, women were on average shorter than men and had a lower mean birthweight (Table 1).

**Table 1:** Characteristics of the sample from the MRC National Survey of Health and Development with complete data on the SITAR height parameters and hip modes (maximum n=769; N varies due to missing data).

|  |  | Men |  |  | Women |  |  |
| --- | --- | --- | --- | --- | --- | --- | --- |
|  |  | N | Mean | SD | N | Mean | SD |
| Height (cm) at age (y): |  |  |  |  |  |  |  |
|  | 2 | 638 | 86.0 | 5.0 | 684 | 85.0 | 4.4 |
|  | 4 | 690 | 103.7 | 4.8 | 752 | 103.5 | 4.9 |
|  | 6 | 658 | 114.8 | 5.0 | 716 | 114.4 | 5.2 |
|  | 7 | 673 | 120.7 | 5.2 | 743 | 120.4 | 5.4 |
|  | 11 | 659 | 141.2 | 6.6 | 722 | 142.0 | 6.9 |
|  | 15 | 616 | 162.6 | 8.9 | 666 | 159.4 | 6.0 |
|  | 20 | 639 | 177.3 | 6.5 | 728 | 163.1 | 6.1 |
| Birthweight (kg) |  | 769 | 3.5 | 0.5 | 838 | 3.4 | 0.5 |
| Weight (kg) at age (y): |  |  |  |  |  |  |  |
|  | 2 | 659 | 13.2 | 1.4 | 714 | 12.7 | 1.5 |
|  | 4 | 713 | 17.6 | 2.1 | 769 | 17.2 | 2.1 |
|  | 6 | 663 | 21.0 | 2.5 | 724 | 20.5 | 2.6 |
|  | 7 | 657 | 23.1 | 2.8 | 718 | 22.8 | 3.1 |
|  | 11 | 659 | 34.4 | 5.8 | 716 | 35.3 | 6.8 |
|  | 15 | 618 | 51.2 | 9.2 | 665 | 52.3 | 8.1 |
|  | 20 | 642 | 70.4 | 8.7 | 723 | 57.8 | 8.2 |
|  |  | N | % |  | N | % |  |
| Sporting ability at 13 years: | Above average | 141 | 20.9 |  | 131 | 18.0 |  |
|  | Average | 417 | 61.8 |  | 527 | 72.5 |  |
|  | Below average | 117 | 17.3 |  | 69 | 9.5 |  |
| Father's occupational class in childhood: | Manual | 360 | 48.7 |  | 400 | 49.7 |  |
|  | Non-manual | 379 | 51.3 |  | 405 | 50.3 |  |

Overall, there was evidence that variations in the patterns of pubertal growth and in height at different ages across childhood and adolescence were associated with hip mode scores at age 60-64 years, with most associations maintained when adjusted for life factors and weight (Tables 1-3 & Suppl. Tables 2 – 4). However, not all associations were found to be equally strong and there was some evidence of deviations from linearity when quadratic terms were included in our models (Table S1). When plots of the associations between height and hip mode were examined in those cases where the quadratic term was statistically significant, this suggested that any deviations from linearity were minor and due to extreme values (Fig. S1).

In sex and early life factor adjusted models, taller height at all ages was associated with positive HM5 & HM6, and negative HM9 scores (Fig. 1 & Table S2). In all other hip modes, the strongest associations varied by age of height assessment, and these tended to be for measures of height in adolescence rather than childhood (Fig. 1 & Table S1). We next examined the SITAR parameters of height size, tempo and velocity with each hip mode (Table 2). When adjusted for sex and early life factors, greater height size was associated with negative mean scores for HM1, HM9 & HM10, and positive scores for HM6 & HM8. In models of HM2 there was evidence of interactions between sex and both height size (p=0.007) and height velocity (p=0.007). In both sexes associations were negative but there was evidence that the associations were stronger in men than women. Greater height velocity was associated with positive mean scores for HM6 and HM7, and negative mean scores for HM10. Later height tempo was associated with negative mean scores for HM2, however there were no associations with any of the other hip modes.

**Figure 1.**
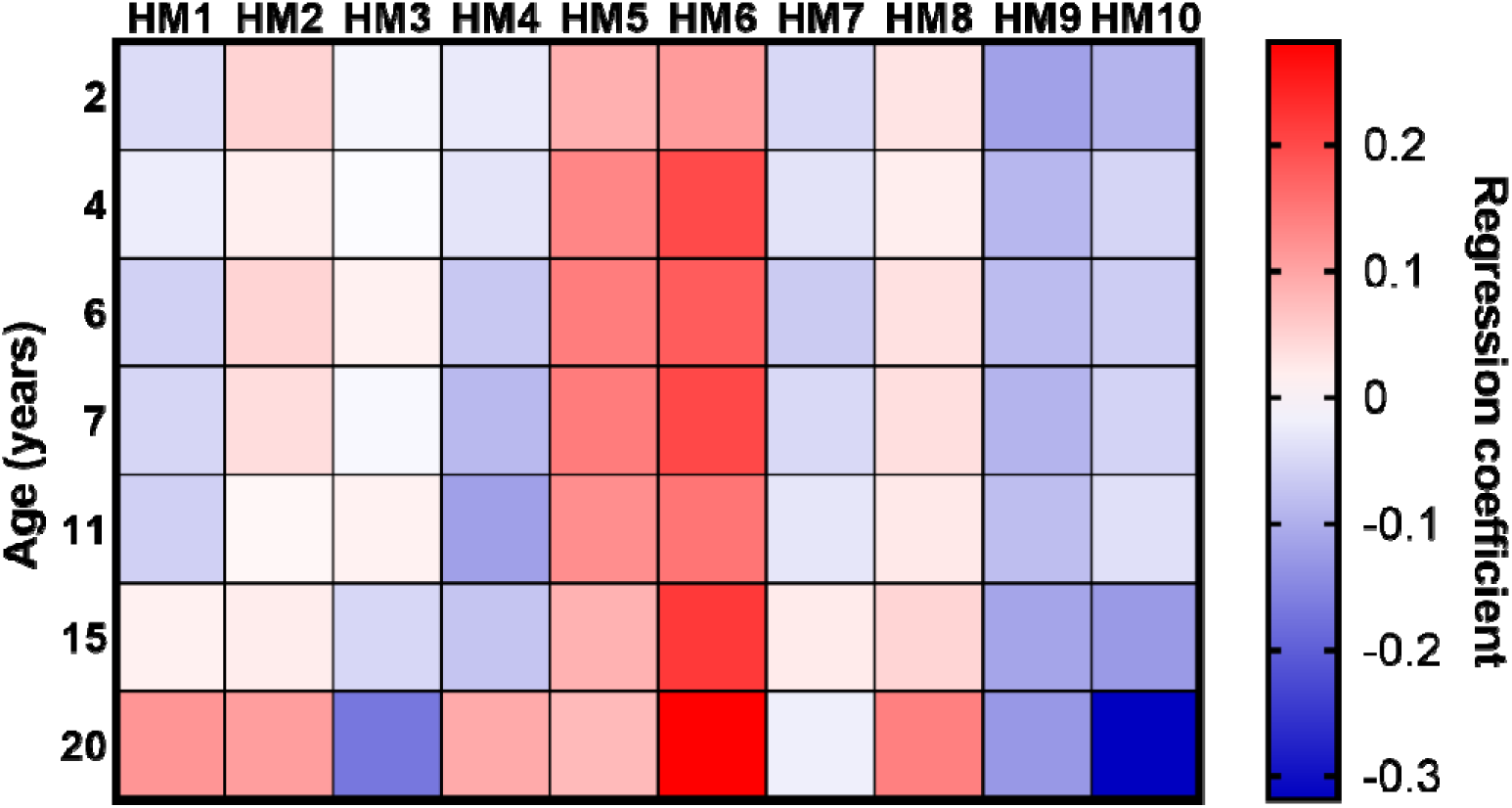
Heat map of the associations between height at ages 2 to 20 years, and each hip shape mode (HM1-HM10). Drawn from the regression coefficients for associations between height assessed at ages 2, 4, 6, 7, 11, 15 and 20 and each hip shape mode score.

**Table 2:**
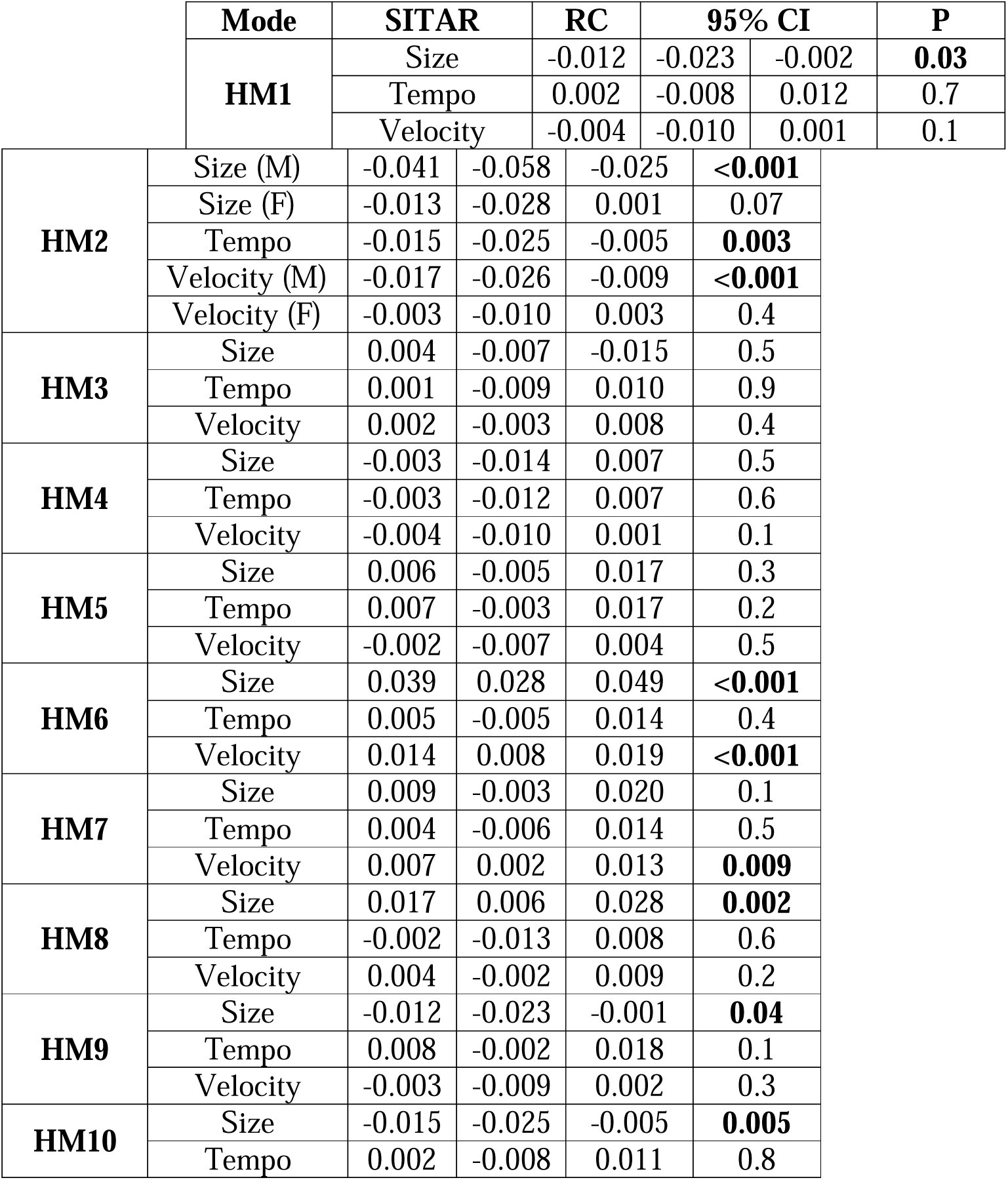
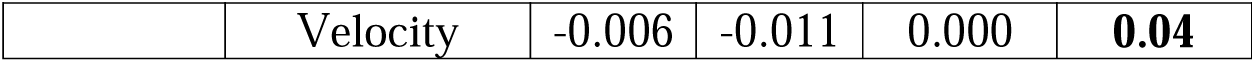
Associations between each parameter of the SITAR model of growth curve analysis (per 1 unit change in height size (cm), tempo (years) and velocity (%)) and HM1-10. All models were run on the sample with valid data for each hip shape mode, each SITAR variable and the confounders. Data presented are from linear regression models adjusted for sex (unless sex interactions were observed), birth weight, sporting ability and father’s occupational class in childhood, as well as SITAR weight. N=1380 (667 male and 713 female). Data from Models 1 & 2 are presented in Table S3.

When examining height gains between specific growth periods, no patterns were consistently found. Where there was any suggestion of an association, this was typically found in relation to changes in height in the 7-15 year period, suggesting these conditional height gains in adolescence rather than childhood may be more important in defining hip shape (HM2 (P<0.001), HM5 (P=0.03), HM6 (P=0.05) & HM7 (Male only – P<0.001); Table 3).

**Table 3:**
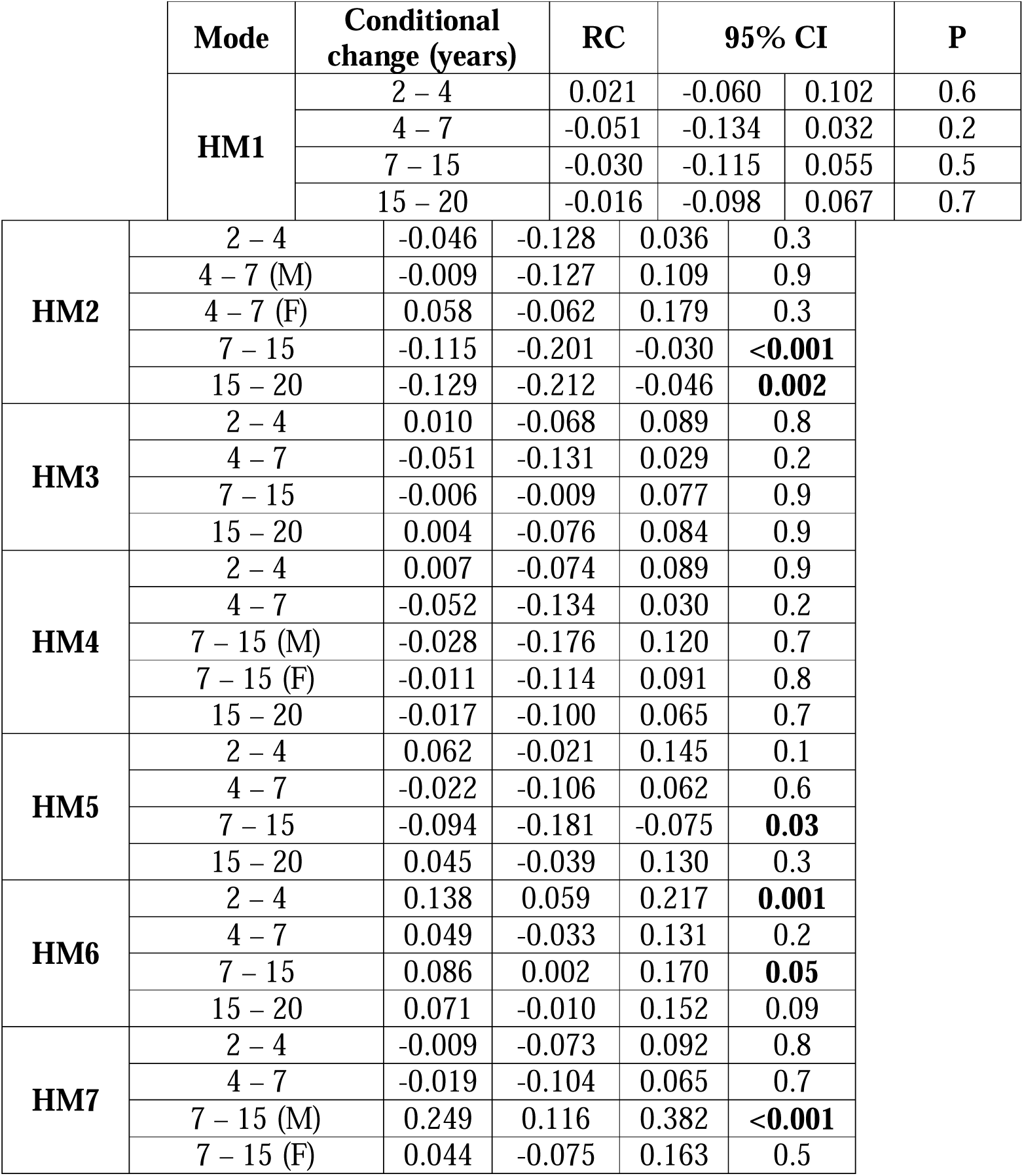
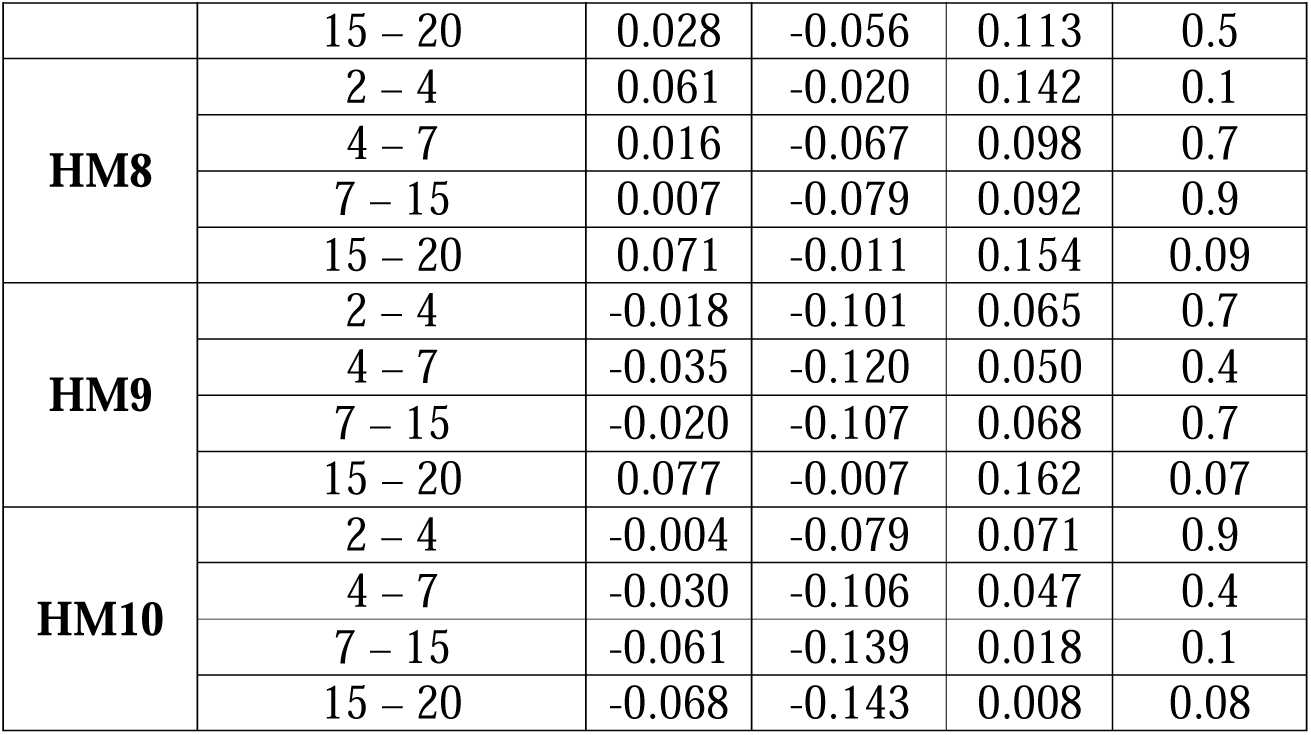
Associations of conditional height gain (per standard deviation) during different periods of growth (early childhood: 2–4 years; late childhood: 4-7 years; childhood to adolescence: 7–15 years; adolescence to young adulthood: 15–20 years) with each hip shape mode. All models were run on the sample with complete data for each hip shape mode, height at each age, and all confounders. Data presented are from linear regression models adjusted for sex (unless sex interactions were observed), birth weight, sporting ability and father’s occupational class in childhood, as well as conditional weight gain. n = 648 (319 male and 329 female).

Subsequent formal comparisons of the 7-15 year period compared to other periods showed evidence that the association at 7 – 15 years was different from the association in all other periods with regards to HM5 and HM7 (P<0.05), with the size of the effect estimates suggesting a stronger association at 7-15 years. However, with regards to HM6, the size of the effect estimates suggesting a stronger association at 2 – 4 years than at 7 – 15 years (P=0.03).

When taking the SITAR findings together, in individuals who are taller, have later pubertal timing (later height tempo), or faster growth velocity, the modes with the most frequent associations (HM2, HM6, and HM10) describe features more consistent with those seen in osteoarthritis, with a wider and flatter femoral head and neck, increased external rotation and presence of osteophytes (Fig. 2). Consistent with these associations with pubertal timing and faster pubertal growth, the majority of associations with hip shape modes were with conditional height gains in adolescence rather than childhood, and greater gains were also associated with osteoarthritis like features. The shape variations described by the associations with each of the HM are shown in Supplementary Table 5.

**Figure 2.**
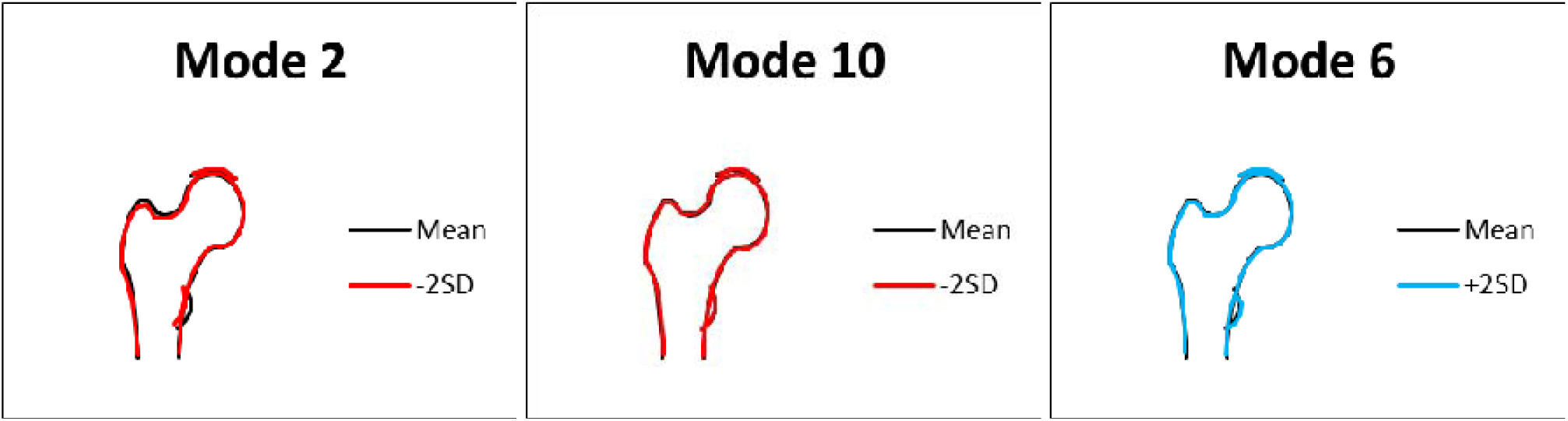
Variations in the modes with the most frequent associations with the SITAR model of growth curve analysis (height size, tempo and velocity) and conditional height gains. Variations in shape for +2 (red) and −2 (blue) SDs in mode score from the mean of 0 for modes 2 and 10 (negative), and 6 (positive). Descriptions of the key features identified by each mode are given in Table S5.

## Discussion

In this study, we aimed to examine associations between longitudinal growth during childhood and adolescence and hip shape at age 60-64 years, assessed by SSM from regional DXA scans in the MRC NSHD British birth cohort. Overall, there was evidence that variations in the patterns of pubertal growth and in height at different ages across childhood and adolescence were modestly associated with hip mode scores at age 60-64 years. Associations were maintained after adjustment for early life factors as well as weight. Examination of height gains during specific periods of childhood and adolescence identified the majority of associations to be in the adolescent period, rather than in childhood.

In both sexes, we observed that faster growth velocity (SITAR height velocity) was associated with changes in hip modes with features more consistent with those seen in osteoarthritis, such as a wider and flatter femoral head and neck, increased external rotation and presence of osteophytes. Similar associations were observed with greater SITAR height size and later height tempo (age at peak height velocity). These features in the hip shapes are similar to those found to be associated with osteoarthritis development in later life in several other studies.

In the Rotterdam Study, a cohort of individuals with no signs of radiographic hip osteoarthritis at baseline and using an active shape model, associations between shape changes in the femoral head and neck, and osteoarthritis onset were examined. The study found that significant changes in the shape of the proximal femur occurred within those individuals who developed osteoarthritis [3]. Similarly, in the Johnston County Osteoarthritis Project, variations with hip shape were associated with prevalent knee radiographic osteoarthritis [19], and in a high bone mass cohort, there was strong evidence of an association between cam morphology (bulging of the lateral femoral head) and radiographic hip osteoarthritis, consistent with other studies [20]. However, there was no evidence of an association between high bone mass and cam morphology, suggestive of distinct pathways by which the risk of osteoarthritis is conferred by these [20]. Therefore, despite the majority of osteoarthritis research focussing on the impact on articular cartilage, these existing studies together with our novel data suggest that bone related changes in shape should be investigated further, especially in the early stages of disease and when considering disease predisposition. It is however important to note that in the current study we examined hip shape at ages 60 – 64 years when the cohort are arguably still relatively young and osteoarthritis prevalence was low, and it would therefore be of interest to further examine these potential associations in this cohort at a later date as they continue to age.

The most obvious mechanism by which hip shape may predict osteoarthritis predisposition is biomechanical, as the shape of the hip may influence joint loading and forces generated. Indeed, in preclinical models, loading through the joint drives significant lasting changes in tibial shape [21]. Alternatively, there may be other genetic and molecular mechanisms which many contribute to this relationship. Several known osteoarthritis susceptibility SNPs have been associated with hip shape in perimenopausal women in ALSPAC, and when combined with data from other cohorts, these SNPs were within 200 kb of genes (*SOX9*, *PTHrP*, *RUNX1*, *NKX3*lJ*2*, *FGFR4*, *DICER1*, *HHIP*) known to be involved in endochondral bone formation [6,7]. A necessary line of research is therefore to evaluate the interaction of endochondral ossification, and in particular these identified genes in determining joint health in later life.

Endochondral ossification is responsible for the development of long bones and occurs at the growth plate throughout childhood and adolescence [22]. It is now well established that in osteoarthritis, the chondrocytes in the articular cartilage revert from their stable phenotype to a more transient one similar to that observed in endochondral ossification [23,24]. Similarly, our previous work exploring associations between growth dynamics and osteoarthritis onset in a spontaneous murine model of osteoarthritis, the STR/Ort mouse revealed accelerated long bone growth, aberrant expression of growth plate markers and enriched growth plate bridging, indicative of advanced and thus premature growth cessation, in these osteoarthritis-prone mice [25]. Together this suggested that these accelerated growth dynamics in young osteoarthritis-prone mice may underpin their osteoarthritis onset and contributes to a growing body of evidence suggestive of an association between longitudinal growth and musculoskeletal health with ageing.

The SITAR growth curve model offers a unique opportunity to effectively examine pubertal growth based on three parameters of size, tempo, and velocity in human cohorts [13,14]. We have previously shown that in the MRC NSHD, there was limited evidence to suggest that height in childhood, as modelled using the SITAR parameters, is associated with odds of knee osteoarthritis in midlife [9]. Our results presented herein similarly suggest that whilst associations between the SITAR parameters and hip shape are modest, individuals who are taller, have later pubertal timing (later height tempo), and/or faster growth velocity have features including a wider and flatter femoral head and neck, increased external rotation and presence of osteophytes. SexLJspecific variation was only identified for associations with HM2 (SITAR height size and velocity, and conditional height gain age 4-7 years), HM4 and HM7 (conditional height gain age 7-15 years). In all cases, associations were stronger in males, consistent with the studies examining HM by Frysz et al., and Ireland et al., and with previous studies of physical activity and bone [26]. These sex differences could reflect a differential response to loading, or potential hormonal effects.

A recent study by Faber et al., conducted the largest GWAS of hip minimum joint space width to date and using cluster analysis identified three distinct clusters [27]. Of these, SNPs in cluster 2 (n = 10) which were associated with a higher osteoarthritis risk with increasing minimum joint space width, showed positive associations with height, and subsequent Mendelian randomisation further showed a strong causal effect of these SNPs on height [27]. This is consistent with previous studies examining the relationship between height and osteoarthritis, and our findings here using the SITAR parameters of height size [28–30]. Faber et al., conclude that as height itself does not appear to be on the causal pathway for minimum joint space width, there is a potential role for an intermediary growth-related mechanism [27]. This further highlights the need for understanding the contribution of endochondral ossification in osteoarthritis predisposition.

Analyses of ALSPAC data have found height tempo to be associated with many of the top hip shape modes at age 14 years, with associations stronger in male participants than female [8]. However, the authors found little evidence of relationships between height tempo and femoral shape measured at age 18 [8]. This is consistent with our studies as herein we found the most consistent pattern of associations with hip shape in later life in the adolescent period of 7-15 years. It has been suggested by Bass et al., that during periods of fast growth specific bone regions may be more responsive to genetic and/or environmental stimuli [31]. Consistent with this, we have previously shown that an early age at onset of walking in infancy is associated with hip shape features which represent an osteoarthritis-like phenotype and suggest that this may relate directly to skeletal loading during a period where skeletal growth is more rapid than at any other time point [10]. Similarly, there are several studies suggesting that high levels of physical activity during another key period of fast growth ie. puberty, may have undesirable effects on the hip shape including cam morphology [32]. Further these studies suggest that these cam-deformities are persistent beyond growth plate closure [33,34]. Further studies are required to fully define the association and underlying mechanisms between growth during these periods and hip shape deformities.

### Strengths and limitations

In this cohort at 60-64 years, the incidence of osteoarthritis was low with the majority of participants having a Kellgren-Lawrence grading of 0 [35]. This may represent selection bias with individuals attending a clinic for DXA assessment more likely to be in better health than those who were visited at home [36]. Another potential limitation is that cross comparison of individual HMs between studies is difficult due to variation in the different models applied and the sensitivity of the models to the data they draw on. Therefore, whilst correlations between our results found here and others can be made as the variation in each HM score can be related to specific features of variation, any conclusive comparisons are difficult. Finally, as with all observational studies, causality cannot be assumed, there is a possibility of residual confounding and results found may be due to change given multiple testing. The major strength of this study is the availability of multiple height measurements prospectively assessed during childhood and adolescence, as well as hip shape SSM modes in a relatively large sample of people at age 60-64 years. Together this provided a unique opportunity to study the relationships between longitudinal growth and hip shape, through two complementary methods: using 1) the SITAR growth curve model to summarise variables of height size, tempo, and velocity 2) a conditional change approach to determine sensitive periods of growth which are associated with joint shape.

### Conclusions

In this relatively large population-based cohort study we have found some evidence to suggest that growth patterns in childhood and adolescence may be associated with modest variations in hip shape at ages 60-64 years, which are consistent with features seen in osteoarthritis. This work contributes to a growing body of work examining how life course height may predict musculoskeletal health in ageing. Further studies are required to fully understand the how sensitive periods of growth during childhood and adolescence may relate to joint shape and ultimately health in ageing.

## Data Availability

All data produced in the present study are available upon reasonable request to the authors

## Acknowledgements

The authors thank all the participants of the MRC National Survey of Health and Development and all staff involved in data collection and data entry, as well as Prof. Tim Cole (SITAR growth curve analysis). Data used in this publication are available to bona fide researchers upon request to the NSHD Data Sharing Committee via a standard application procedure. Further details can be found at http://www.nshd.mrc.ac.uk/data. doi: 10.5522/NSHD/Q101 and 10.5522/NSHD/S102A.

## Author contributions

All authors contributed to the conception and design of the study, or acquisition of data, or analysis and interpretation of data; drafting the article or revising it critically for important intellectual content and the final approval of the version to be submitted. KS takes responsibility for the integrity of the work as a whole, from inception to finished article.

