## Supplementary material for "Associations between life course longitudinal growth and hip shapes at ages 60 to 64 years: evidence from the MRC National Survey of Health and Development": Suppl data

**Supplementary data: Associations between life course longitudinal growth and hip shapes at age 60 to 64: evidence from the 1946 British birth cohort study**

Katherine A. Staines., Fiona Saunders., Alex Ireland., Richard Aspden., Jenny Gregory., Rebecca Hardy., Rachel Cooper

**Table S1.** *Unadjusted differences in height (per 5cm) and SITAR parameters (per 1 unit change in height size (cm), and velocity (%)) with hip shape mode, where there was evidence of deviations from linearity when a quadratic term was included in the main model.*

| **Mode** | **Height / 5cm at age** |  | **RC** | **95% CI** | | **P** |
| --- | --- | --- | --- | --- | --- | --- |
| HM8 | 7 | Linear | 0.424 | -0.096 | 0.752 | 0.01 |
|  |  | Quadratic | -0.002 | -0.003 | -0.000 | 0.02 |
| HM9 | 6 | Linear | 0.371 | 0.033 | 0.709 | 0.03 |
|  |  | Quadratic | -0.002 | -0.003 | -0.000 | 0.03 |
| HM9 | 20 | Linear | 0.255 | 0.071 | 0.438 | 0.007 |
|  |  | Quadratic | -0.009 | -0.001 | -0.000 | 0.005 |
|  | **SITAR** |  |  |  | |  |
| HM9 | Height size | Linear | -0.012 | -0.021 | -0.003 | 0.01 |
|  |  | Quadratic | -0.002 | -0.003 | -0.001 | 0.005 |
| HM10 | Height velocity | Linear | -0.006 | -0.011 | -0.001 | 0.02 |
|  |  | Quadratic | 0.000 | 0.000 | 0.001 | 0.03 |

***Table S2:*** *Associations between height (per 5cm) at different ages throughout childhood, adolescence and young adulthood and each hip shape mode. All models were run on the sample with valid data for hip shape modes, height at the specific age and the confounders, adjusted for sex (unless sex interactions were observed). n = 2 years – 1113; 4 years – 1248; 6 years – 1186; 7 years – 1247; 11 years – 1271; 15 years – 1183; 20 years – 1201.*

| **Mode** | **Height / 5cm at age** | **RC** | **95% CI** | | **P** |
| --- | --- | --- | --- | --- | --- |
| **HM1** | 2 | -0.022 | -0.083 | 0.040 | 0.5 |
|  | 4 | -0.048 | -0.104 | 0.008 | 0.1 |
|  | 6 | -0.087 | -0.143 | -0.031 | **0.002** |
|  | 7 | -0.072 | -0.124 | -0.020 | **0.007** |
|  | 11 | -0.047 | -0.087 | -0.007 | **0.02** |
|  | 15 | -0.022 | -0.059 | 0.015 | 0.3 |
|  | 20 | -0.050 | -0.095 | -0.005 | **0.03** |
| **HM2** | 2 | 0.245 | -0.037 | 0.086 | 0.4 |
|  | 4 | 0.012 | -0.045 | 0.068 | 0.7 |
|  | 6 (M) | -0.029 | -0.115 | 0.057 | 0.5 |
|  | 6 (F) | 0.115 | 0.041 | 0.190 | **0.003** |
|  | 7 (M) | -0.057 | -0.135 | 0.022 | 0.2 |
|  | 7 (F) | 0.079 | 0.009 | 0.149 | **0.03** |
|  | 11 (M) | -0.065 | -0.125 | 0.005 | **0.03** |
|  | 11 (F) | 0.085 | 0.032 | 0.139 | **0.002** |
|  | 15 (M) | -0.032 | -0.078 | 0.014 | 0.2 |
|  | 15 (F) | 0.040 | -0.024 | 0.103 | 0.2 |
|  | 20 (M) | -0.079 | -0.145 | -0.013 | **0.02** |
|  | 20 (F) | 0.022 | -0.039 | 0.084 | 0.5 |
| **HM3** | 2 | -0.029 | -0.089 | 0.031 | 0.3 |
|  | 4 | -0.044 | -0.099 | 0.011 | 0.1 |
|  | 6 | -0.034 | -0.088 | 0.020 | 0.2 |
|  | 7 | -0.057 | -0.107 | -0.064 | **0.03** |
|  | 11 | -0.034 | -0.074 | 0.008 | 0.09 |
|  | 15 | -0.030 | -0.066 | 0.006 | 0.1 |
|  | 20 | -0.015 | -0.058 | 0.028 | 0.5 |
| **HM4** | 2 | -0.042 | -0.102 | 0.019 | 0.2 |
|  | 4 | -0.023 | -0.079 | 0.032 | 0.4 |
|  | 6 | -0.045 | -0.100 | 0.009 | 0.1 |
|  | 7 | -0.061 | -0.113 | -0.100 | **0.02** |
|  | 11 | -0.061 | -0.100 | -0.021 | **0.003** |
|  | 15 | -0.074 | -0.110 | -0.037 | **<0.001** |
|  | 20 | -0.034 | -0.077 | 0.010 | 0.1 |
| **HM5** | 2 | 0.101 | 0.039 | 0.164 | **0.001** |
|  | 4 | 0.107 | 0.050 | 0.164 | **<0.001** |
|  | 6 | 0.108 | 0.052 | 0.165 | **<0.001** |
|  | 7 | 0.093 | 0.040 | 0.146 | **0.001** |
|  | 11 | 0.077 | 0.037 | 0.118 | **<0.001** |
|  | 15 | 0.039 | 0.002 | 0.077 | **0.04** |
|  | 20 | 0.066 | 0.021 | 0.116 | **0.004** |
| **HM6** | 2 | 0.116 | 0.057 | 0.176 | **<0.001** |
|  | 4 | 0.165 | 0.111 | 0.220 | **<0.001** |
|  | 6 | 0.148 | 0.094 | 0.202 | **<0.001** |
|  | 7 | 0.156 | 0.105 | 0.207 | **<0.001** |
|  | 11 | 0.112 | 0.072 | 0.151 | **<0.001** |
|  | 15 | 0.110 | 0.074 | 0.146 | **<0.001** |
|  | 20 | 0.161 | 0.118 | 0.205 | **<0.001** |
| **HM7** | 2 | -0.048 | -0.111 | 0.015 | 0.1 |
|  | 4 | -0.041 | -0.098 | 0.016 | 0.2 |
|  | 6 | -0.068 | -0.124 | -0.012 | **0.02** |
|  | 7 | -0.040 | -0.093 | 0.013 | 0.1 |
|  | 11 | -0.022 | -0.063 | 0.018 | 0.3 |
|  | 15 | 0.016 | -0.021 | 0.054 | 0.4 |
|  | 20 | -0.001 | -0.045 | 0.044 | 1.0 |
| **HM8** | 2 | 0.061 | -0.001 | 0.123 | **0.05** |
|  | 4 | 0.025 | -0.032 | 0.081 | 0.4 |
|  | 6 | 0.031 | -0.025 | 0.086 | 0.3 |
|  | 7 | 0.036 | -0.016 | 0.088 | 0.2 |
|  | 11 | 0.043 | 0.003 | 0.083 | **0.03** |
|  | 15 | 0.026 | -0.010 | 0.062 | 0.2 |
|  | 20 | 0.049 | 0.004 | 0.093 | **0.03** |
| **HM9** | 2 | -0.117 | -0.179 | -0.055 | **<0.001** |
|  | 4 | -0.071 | -0.127 | -0.015 | **0.01** |
|  | 6 | -0.081 | -0.137 | -0.025 | **0.004** |
|  | 7 | -0.102 | -0.155 | -0.049 | **<0.001** |
|  | 11 | -0.083 | -0.123 | -0.042 | **<0.001** |
|  | 15 | -0.064 | -0.101 | -0.026 | **0.001** |
|  | 20 | -0.043 | -0.088 | 0.002 | 0.06 |
| **HM10** | 2 | -0.034 | -0.093 | 0.023 | 0.2 |
|  | 4 | -0.037 | -0.090 | 0.017 | 0.2 |
|  | 6 | -0.009 | -0.062 | 0.044 | 0.7 |
|  | 7 | -0.045 | -0.095 | 0.005 | 0.8 |
|  | 11 | -0.037 | -0.075 | 0.001 | 0.06 |
|  | 15 | -0.035 | -0.067 | 0.000 | **0.05** |
|  | 20 | -0.065 | -0.108 | -0.023 | **0.002** |

***Table S3:*** *Associations between each parameter of the SITAR model of growth curve analysis (per 1 unit change in height size (cm), tempo (years) and velocity (%)) and HM1-10. All models were run on the sample with valid data for each hip shape mode, each SITAR variable and the confounders. Data presented are from linear regression models adjusted for Model 1: sex (unless sex interactions were observed); Model 2: birth weight, sporting ability and Father’s occupational class in childhood. n=1380 (667 male and 713 female).*

|  |  | **Model 1** | | | | **Model 2** | | | |
| --- | --- | --- | --- | --- | --- | --- | --- | --- | --- |
| **Mode** | **SITAR** | **RC** | **95% CI** | | **P** | **RC** | **95% CI** | | **P** |
| **HM1** | Size | -0.010 | -0.019 | -0.001 | **0.03** | -0.012 | -0.021 | -0.002 | **0.015** |
|  | Tempo | 0.003 | -0.005 | 0.010 | 0.5 | 0.002 | -0.006 | 0.010 | 0.6 |
|  | Velocity | -0.003 | -0.008 | 0.002 | 0.2 | -0.004 | -0.009 | 0.001 | 0.2 |
| **HM2** | Size (M) | -0.020 | -0.034 | -0.007 | **0.03** | -0.021 | -0.035 | -0.007 | **0.003** |
|  | Size (F) | 0.004 | -0.008 | 0.017 | 0.5 | 0.004 | -0.009 | 0.017 | 0.6 |
|  | Tempo | -0.011 | -0.019 | -0.003 | **0.005** | -0.011 | -0.019 | -0.004 | **0.004** |
|  | Velocity (M) | -0.012 | -0.020 | -0.004 | **0.002** | -0.012 | -0.020 | -0.004 | **0.002** |
|  | Velocity (F) | 0.002 | -0.005 | 0.009 | 0.6 | 0.002 | -0.005 | 0.009 | 0.6 |
| **HM3** | Size | -0.004 | -0.013 | 0.005 | 0.4 | -0.001 | -0.010 | 0.008 | 0.8 |
|  | Tempo | 0.005 | -0.003 | 0.013 | 0.2 | 0.005 | -0.003 | 0.012 | 0.2 |
|  | Velocity | 0.001 | -0.004 | 0.006 | 0.8 | 0.001 | -0.004 | 0.006 | 0.6 |
| **HM4** | Size | -0.011 | -0.019 | -0.002 | **0.02** | -0.009 | -0.019 | -0.000 | **0.05** |
|  | Tempo | 0.007 | -0.001 | 0.015 | 0.07 | 0.007 | -0.001 | 0.015 | 0.07 |
|  | Velocity | -0.006 | -0.011 | -0.001 | **0.02** | -0.006 | -0.011 | -0.007 | **0.03** |
| **HM5** | Size | 0.015 | 0.005 | 0.024 | **0.002** | 0.014 | 0.004 | 0.023 | **0.006** |
|  | Tempo | -0.003 | -0.011 | 0.005 | 0.5 | -0.002 | -0.010 | 0.006 | 0.6 |
|  | Velocity | 0.002 | -0.004 | 0.007 | 0.5 | 0.001 | -0.004 | 0.007 | 0.6 |
| **HM6** | Size | 0.038 | 0.029 | 0.046 | **<0.001** | 0.039 | 0.030 | 0.048 | **<0.001** |
|  | Tempo | 0.003 | -0.005 | 0.011 | 0.5 | 0.003 | -0.005 | 0.010 | 0.5 |
|  | Velocity | 0.015 | 0.010 | 0.020 | **<0.001** | 0.015 | 0.010 | 0.020 | **<0.001** |
| **HM7** | Size | 0.000 | -0.009 | 0.009 | 1.0 | 0.001 | -0.010 | 0.010 | 0.9 |
|  | Tempo | 0.002 | -0.006 | 0.001 | 0.6 | 0.002 | -0.006 | 0.010 | 0.6 |
|  | Velocity | 0.005 | -0.000 | 0.101 | 0.07 | 0.005 | -0.000 | 0.103 | **0.06** |
| **HM8** | Size | 0.011 | 0.002 | 0.020 | **0.02** | 0.011 | 0.002 | 0.021 | **0.02** |
|  | Tempo | 0.002 | -0.005 | 0.101 | 0.6 | 0.002 | -0.005 | 0.010 | 0.5 |
|  | Velocity | 0.003 | -0.003 | 0.008 | 0.3 | 0.003 | -0.003 | 0.008 | 0.3 |
| **HM9** | Size | -0.013 | -0.022 | -0.004 | **0.005** | -0.014 | 0.023 | -0.004 | **0.005** |
|  | Tempo | 0.010 | 0.002 | 0.018 | **0.01** | 0.010 | 0.003 | 0.018 | **0.01** |
|  | Velocity | -0.004 | -0.009 | 0.001 | 0.1 | -0.004 | -0.009 | 0.001 | 0.2 |
| **HM10** | Size | -0.013 | -0.022 | -0.004 | **0.003** | -0.013 | -0.022 | -0.004 | **0.005** |
|  | Tempo | -0.002 | -0.009 | 0.006 | 0.7 | -0.002 | -0.009 | 0.006 | 0.6 |
|  | Velocity | -0.006 | -0.011 | -0.001 | **0.03** | -0.006 | -0.010 | -0.001 | **0.03** |

***Table S4:*** *Associations of conditional height gain (per standard deviation) during different periods of growth (early childhood: 2–4 years; childhood to adolescence: 7–15 years; adolescence to young adulthood: 15–20 years) with each hip shape mode. All models were run on the sample with complete data for each hip shape mode, height at each age, and the confounders. Data presented are from linear regression models adjusted for Model 1: sex (unless sex interactions were observed); Model 2: birth weight, sporting ability and Father’s occupational class in childhood.* *n = 648 (319 male and 329 female).*

|  |  | Model 1 | | | | Model 2 | | | |
| --- | --- | --- | --- | --- | --- | --- | --- | --- | --- |
| Mode | Conditional change (years) | RC | 95% CI | | P | RC | 95% CI | | P |
| **HM1** | 2 – 4 | -0.001 | -0.078 | 0.076 | 1.0 | -0.003 | -0.080 | 0.074 | 0.9 |
|  | 4 – 7 | -0.049 | -0.127 | 0.028 | 0.2 | -0.052 | -0.128 | 0.024 | 0.2 |
|  | 7 – 15 | 0.014 | -0.063 | 0.091 | 0.7 | -0.008 | -0.068 | 0.084 | 0.8 |
|  | 15 – 20 | -0.017 | -0.093 | 0.058 | 0.7 | 0.006 | -0.081 | 0.069 | 0.9 |
| **HM2** | 2 – 4 | -0.011 | -0.087 | 0.065 | 0.8 | -0.012 | -0.089 | 0.067 | 0.8 |
|  | 4 – 7 (M) | -0.019 | -0.127 | 0.089 | 0.7 | -0.021 | -0.11 | 0.088 | 0.7 |
|  | 4 – 7 (F) | 0.071 | -0.039 | 0.181 | 0.2 | -0.047 | -0.124 | 0.029 | 0.2 |
|  | 7 – 15 | -0.046 | -0.122 | 0.030 | 0.2 | -0.041 | -0.102 | 0.021 | 0.2 |
|  | 15 – 20 | -0.099 | -0.174 | -0.024 | **0.01** | -0.097 | -0.172 | -0.022 | **0.01** |
| **HM3** | 2 – 4 | 0.005 | -0.069 | 0.078 | 0.9 | -0.006 | -0.069 | 0.080 | 0.9 |
|  | 4 – 7 | -0.040 | -0.113 | 0.034 | 0.3 | -0.044 | -0.118 | 0.030 | 0.2 |
|  | 7 – 15 | -0.012 | -0.085 | 0.061 | 0.7 | 0.004 | -0.078 | 0.069 | 0.9 |
|  | 15 – 20 | -0.009 | -0.082 | 0.063 | 0.8 | -0.005 | -0.078 | 0.067 | 0.9 |
| **HM4** | 2 – 4 | -0.016 | -0.091 | 0.059 | 0.7 | 0.002 | -0.075 | 0.079 | 1.0 |
|  | 4 – 7 | -0.078 | -0.154 | -0.02 | **0.04** | -0.071 | -0.147 | 0.05 | 0.1 |
|  | 7 – 15 (M) | -0.098 | -0.214 | 0.018 | 0.1 | -0.095 | -0.212 | 0.022 | 0.1 |
|  | 7 – 15 (F) | -0.011 | -0.107 | 0.084 | 0.8 | -0.053 | -0.103 | 0.092 | 0.9 |
|  | 15 – 20 | -0.001 | -0.076 | 0.073 | 1.0 | 0.002 | -0.073 | 0.077 | 1.0 |
| **HM5** | 2 – 4 | 0.090 | 0.014 | 0.167 | **0.02** | 0.080 | 0.002 | 0.159 | **0.04** |
|  | 4 – 7 | 0.045 | -0.033 | 0.123 | 0.3 | 0.041 | -0.037 | 0.119 | 0.3 |
|  | 7 – 15 | -0.057 | -0.134 | 0.020 | 0.1 | -0.061 | -0.138 | 0.017 | 0.1 |
|  | 15 – 20 | 0.052 | -0.024 | 0.129 | 0.2 | 0.043 | -0.033 | 0.119 | 0.3 |
| **HM6** | 2 – 4 | 0.170 | 0.096 | 0.243 | **0.0** | 0.165 | 0.090 | 0.240 | **0.0** |
|  | 4 – 7 | 0.048 | -0.027 | 0.123 | 0.2 | 0.042 | -0.033 | 0.118 | 0.3 |
|  | 7 – 15 | 0.064 | -0.101 | 0.138 | 0.1 | 0.063 | -0.012 | 0.138 | 0.1 |
|  | 15 – 20 | 0.800 | 0.007 | 0.153 | **0.03** | 0.078 | 0.005 | 0.152 | **0.04** |
| **HM7** | 2 – 4 | -0.041 | -0.118 | 0.036 | 0.3 | -0.027 | -0.106 | 0.051 | 0.5 |
|  | 4 – 7 | -0.039 | -0.117 | 0.039 | 0.3 | -0.032 | -0.110 | 0.046 | 0.4 |
|  | 7 – 15 (M) | 0.171 | 0.067 | 0.276 | **0.001** | 0.168 | 0.062 | 0.274 | **0.002** |
|  | 7 – 15 (F) | 0.015 | -0.097 | 0.126 | 0.8 | 0.022 | -0.092 | 0.135 | 0.7 |
|  | 15 – 20 | -0.070 | -0.083 | 0.069 | 0.9 | 0.000 | -0.076 | 0.076 | 1.0 |
| **HM8** | 2 – 4 | 0.024 | -0.051 | 0.100 | 0.5 | 0.037 | -0.040 | 0.114 | 0.4 |
|  | 4 – 7 | 0.013 | -0.063 | 0.089 | 0.7 | 0.016 | -0.060 | 0.092 | 0.7 |
|  | 7 – 15 | -0.017 | -0.092 | 0.058 | 0.7 | -0.011 | -0.087 | 0.065 | 0.8 |
|  | 15 – 20 | 0.062 | -0.012 | 0.136 | 0.1 | 0.066 | -0.009 | 0.140 | 0.08 |
| **HM9** | 2 – 4 | -0.043 | -0.120 | 0.034 | 0.3 | -0.030 | -0.109 | 0.049 | 0.5 |
|  | 4 – 7 | -0.030 | -0.108 | 0.048 | 0.5 | -0.025 | -0.103 | 0.053 | 0.5 |
|  | 7 – 15 | -0.018 | -0.095 | 0.059 | 0.6 | -0.009 | -0.087 | 0.068 | 0.8 |
|  | 15 – 20 | 0.066 | -0.010 | 0.142 | 0.09 | 0.068 | -0.008 | 0.145 | 0.08 |
| **HM10** | 2 – 4 | -0.027 | -0.096 | 0.042 | 0.4 | -0.021 | -0.092 | 0.049 | 0.6 |
|  | 4 – 7 | -0.032 | -0.101 | 0.038 | 0.4 | -0.029 | -0.099 | 0.041 | 0.4 |
|  | 7 – 15 | -0.043 | -0.112 | 0.026 | 0.2 | -0.041 | -0.111 | 0.029 | 0.2 |
|  | 15 – 20 | -0.053 | -0.121 | 0.016 | 0.1 | -0.052 | -0.120 | 0.017 | 0.1 |

***Table S5:*** *Shape variations described by the associations with each of the HM and the SITAR variables and conditional height periods.*

| **Greater SITAR height size** | |
| --- | --- |
| HM1 negative | More compact femoral head  Larger neck-shaft angle |
| HM2 negative (male only) | Longer femoral neck  Increased external rotation  Loss of femoral head curvature |
| HM9 negative | Wider femoral neck  Increasing osteophytes  More compact femoral head |
| HM10 negative | Flatter femoral neck curvature  Medial enlargement of femoral head  Narrower femoral shaft |
| HM6 positive | Some evidence of external rotation from positive to negative scores |
| HM8 positive | Slight medial migration of femoral head  Slightly larger lesser trochanter |
| **Later SITAR height tempo** | |
| HM2 negative | Longer femoral neck  Increased external rotation  Loss of femoral head curvature |
| **Greater SITAR height velocity** | |
| HM2 negative (male only) | Longer femoral neck  Increased external rotation  Loss of femoral head curvature |
| HM10 negative | Flatter femoral neck curvature  Medial enlargement of femoral head  Narrower femoral shaft |
| HM6 positive | Some evidence of external rotation from positive to negative scores |
| HM7 positive | Wider, flatter femoral head  Shorter femoral neck  Slight external rotation |
| **Conditional change 2 – 4 years** | |
| HM6 positive | Some evidence of external rotation from positive to negative scores |
| **Conditional change 7 – 15 years** | |
| HM2 negative | Longer femoral neck  Increased external rotation  Loss of femoral head curvature |
| HM5 negative | Possible external rotation (more of the lesser trochanter visible) |
| HM6 positive | Some evidence of external rotation from positive to negative scores |
| HM7 positive (males only) | Wider, flatter femoral head  Shorter femoral neck  Slight external rotation |
| **Conditional change 15 – 20 years** | |
| HM2 negative | Longer femoral neck  Increased external rotation  Loss of femoral head curvature |

**Figure S1:** *Scatter plots of weighted regression of height (per 5cm) and SITAR parameters with hip shape mode,*

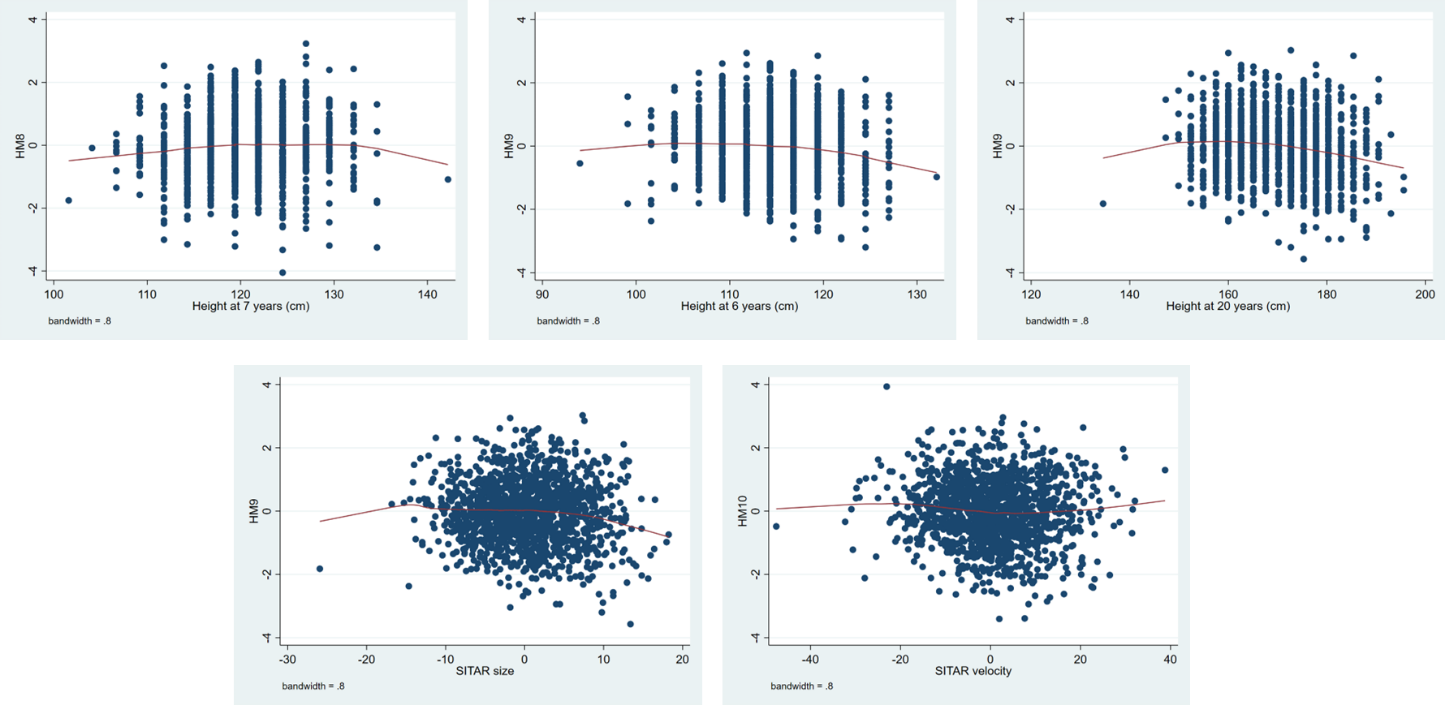
